# Machine Learning Risk Prediction for Prolonged Hospitalization in Frail Older Adults with Multimorbidity

**DOI:** 10.1101/2025.10.18.25338267

**Authors:** Innocent Tesha, Wang Jiasi, Zhao Xizhe, Nassor Makame, Maryam Mbarak, Ding Lin, Yue Chen, Maxwell Ahiafor, Sidney Amadi, Njoka Irene, Jermaine Sikombe, Mwila Kafwembe, Deogratius Galikano, Masoud Mtore, Wellington Ngari, Liu Xinyu

## Abstract

**Background:** Frailty and multimorbidity are common in older adults and contribute substantially to prolonged hospitalizations, readmissions, and mortality. Yet, existing prediction models often fail to integrate frailty-specific biomarkers and lack interpretability for routine clinical use.

**Objectives:** To develop and internally validate an interpretable, machine learning–enhanced logistic regression model to predict prolonged hospital length of stay (LOS) among frail older adults with multimorbidity, and to identify key predictors to guide individualized inpatient care.

**Methods:** We conducted a retrospective study of 440 hospitalized adults aged ≥65 years with multimorbidity (≥2 chronic conditions) and frailty (Frailty Index ≥0.25) at a tertiary geriatric department between January 2022 and December 2023. Fourteen demographic, clinical, and biochemical variables were analysed. Feature selection employed Elastic Net regularization, Extreme Gradient Boosting with SHAP value analysis, and the Boruta algorithm to ensure robust predictor identification. A multivariable logistic regression model was trained and internally validated using stratified 10-fold cross-validation and 1,000 bootstrap iterations. Discrimination (AUC-ROC), calibration, and clinical utility (decision curve analysis) were assessed.

**Results:** Eight predictors age, diabetes, hypertension, prior stroke, serum albumin, HDL cholesterol, systolic blood pressure, and neutrophil-to-lymphocyte ratio—were retained in the final model. The model achieved good discrimination (AUC = 0.770, 95% CI 0.688–0.853) and acceptable calibration (Hosmer–Lemeshow χ² = 14.86, p = 0.062). Cross-validation (mean AUC 0.687 ± 0.072) and bootstrap correction (AUC 0.672) confirmed internal stability. Serum albumin was the strongest protective factor, while elevated neutrophil-to-lymphocyte ratio and prior stroke were significant risk factors.

**Conclusions:** This interpretable model accurately predicts prolonged hospital stay in frail older adults with multimorbidity using routinely available clinical data. Its transparent design supports integration into electronic health records for real-time risk stratification, facilitating targeted discharge planning and personalized geriatric care.

## Introduction

The progressive decline in physiological reserve and functional capacity associated with aging has contributed to a rapid increase in the prevalence of multimorbidity among older adults worldwide. Multimorbidity, defined as the coexistence of two or more chronic conditions [1], affects more than one-third of adults globally, with individuals aged 60 years and older representing over half of this group [2]. The clustering of chronic diseases such as cardiovascular disorders, diabetes mellitus, chronic respiratory diseases, and cancer not only increases healthcare utilization but also substantially elevates the risk of hospital readmission, prolonged hospitalization, and mortality. The interaction between these conditions may exacerbate the cumulative disease burden, thereby significantly compromising patients’ functional capacity, quality of life and overall health[3]. Collectively, these conditions account for more than 80% of premature deaths and remain the dominant contributors to the global burden of noncommunicable diseases[4]

Frailty, a distinct yet overlapping construct with multimorbidity, reflects a state of heightened vulnerability to physiological stressors resulting from cumulative declines across multiple biological systems. It manifests through features such as weakness, slowed mobility, exhaustion, weight loss, and low physical activity[5]. Among hospitalized older adults, frailty has emerged as a powerful independent predictor of adverse outcomes, including functional decline, readmission, and death. Although multimorbidity and frailty are conceptually distinct, they frequently coexist and interact synergistically amplifying risk trajectories and complicating clinical management in geriatric populations [6], [7].

Despite extensive research on the epidemiology of frailty and multimorbidity, predictive tools that accurately estimate clinical outcomes in this vulnerable group remain limited. Conventional frailty instruments such as the Fried phenotype or simple cumulative deficit indices capture physical vulnerability but inadequately represent the complex interplay between inflammation, nutrition, and cardiovascular instability [8].

Similarly, existing risk models in older adults with multimorbidity have primarily focused on single outcomes, such as mortality, and have rarely incorporated dynamic, multidimensional biomarkers. As a result, clinicians often lack reliable, interpretable tools to identify frail older adults at high risk for prolonged hospital stay or delayed recovery, both of which have major implications for quality of care and resource allocation.

Recent advances in machine learning (ML) provide an opportunity to address these gaps. ML algorithms can integrate heterogeneous data demographic, clinical, and biochemical to uncover nonlinear interactions that traditional regression models often overlook. However, the clinical adoption of ML tools in geriatric care has been slow, largely due to concerns about their black-box nature, limited interpretability, and uncertain generalizability [9]. For clinical decision-making in older adults, interpretability and transparency are as crucial as predictive accuracy. Models that combine the explanatory clarity of traditional regression with the feature selection power of ML methods may thus offer a practical and clinically acceptable middle ground [10].

Building on this rationale, the present study aimed to develop and internally validate an interpretable, machine learning–enhanced logistic regression model to predict prolonged hospital length of stay among frail older adults with multimorbidity. By identifying and quantifying the contribution of key demographic, clinical, and laboratory predictors, this study seeks to advance risk stratification strategies in geriatric inpatient care. Ultimately, the model aims to support clinicians in implementing individualized, frailty-informed interventions that improve hospital efficiency and optimize outcomes for older adults with complex health needs.

## Materials and Methods

### Study Design and Ethical Approval

This retrospective observational study was conducted at the Geriatrics Department of the First Affiliated Hospital of Jinzhou Medical University, China. Ethical approval was obtained from the institutional review board (approval number: JZ2023B057). Given the retrospective design, the requirement for informed consent was waived. All procedures adhered to the ethical principles of the Declaration of Helsinki and followed the Transparent Reporting of a Multivariable Prediction Model for Individual Prognosis or Diagnosis (TRIPOD) and TRIPOD-AI guidelines for predictive modelling studies.

### Study Population

Electronic medical records were reviewed for all older adults aged ≥65 years who were hospitalized between January 1, 2022, and December 31, 2023. Inclusion criteria were:

1. Diagnosis of multimorbidity, defined as the coexistence of two or more chronic conditions coded according to ICD-10.
2. Frailty, operationalized as a Frailty Index (FI) ≥0.25, consistent with standard cumulative deficit methodology.
3. Availability of complete demographic, clinical, and laboratory data.

Exclusion criteria included incomplete medical records, frailty index <0.25, active malignancy under palliative care, or terminal illness.

Out of 597 initially screened inpatients, 440 met the inclusion criteria. The sample size was determined using the events per variable (EPV) method for logistic regression [11].

Assuming 14 predictor variables and a frailty prevalence among older adults with multimorbidity of 27.2% as reported by Lv et al [12] a minimum of 414 participants was required to achieve an events-per-variable (EPV) ratio of 8, with an additional 5% attrition accounted for. The final cohort of 440 patients satisfied these sample size requirements.

### Frailty and Multimorbidity Assessment

Frailty was quantified using a 30-item cumulative deficit Frailty Index (FI) constructed in accordance with validated procedures for hospitalized older adults[13]. The FI included items spanning comorbidities, physical function, nutrition, cognition, and psychosocial status (see Supplementary Table 1). Each deficit was scored as 0 (absent) or 1 (present), and the index was calculated as the ratio of present deficits to total possible deficits. A threshold of FI ≥ 0.25 denoted frailty [14].

Multimorbidity was defined as the coexistence of at least two chronic diseases, based on ICD-10 codes. The following 12 chronic conditions were included: coronary heart disease, chronic heart failure, cerebrovascular disease, chronic kidney disease, chronic liver disease, chronic obstructive pulmonary disease, diabetes mellitus, hypertension, cancer, depression, bone or joint disease, and chronic gastrointestinal disease [15].

### Predictor Variables and Clinical Outcomes

Candidate predictors included demographic, clinical, and biochemical parameters routinely collected at hospital admission. Demographic variables: age, sex, smoking status, Clinical history: hypertension, diabetes mellitus, prior stroke. Vital and laboratory markers: systolic blood pressure, haemoglobin, neutrophil count, neutrophil-to-lymphocyte ratio (NLR), serum albumin, creatinine, total cholesterol, HDL cholesterol, and non-HDL cholesterol. The primary outcome was prolonged hospital length of stay (LOS), defined as hospitalization exceeding the third quartile (>8 days) of the cohort’s LOS distribution.

### Data Pre-processing and Feature Selection

Missing data across all variables ranged from 2% to 18%. To minimize bias, missing values were imputed using multivariate imputation by chained equations (MICE), generating complete datasets for subsequent analyses. Continuous variables were standardized using z-score normalization, and categorical variables were encoded as binary indicators. A multi-algorithmic feature selection ensemble was implemented to enhance robustness and interpretability. Elastic Net Regularization was applied with 10-fold cross-validation to optimize the alpha (mixing) and lambda (penalty) parameters, balancing feature sparsity and stability [16]. Extreme Gradient Boosting (XGBoost) with SHAP (SHapley Additive exPlanations) values quantified feature importance across gain, cover, and frequency metrics, with early stopping after 10 rounds to prevent overfitting [17]. Boruta algorithm, a Random Forest–based wrapper, identified all-relevant features by comparing predictor importance against randomized shadow variables across 100 iterations [18]. To synthesize these diverse approaches, Normalized importance scores from all three methods were combined using an ensemble ranking system, and predictors consistently identified across algorithms were retained for model development

### Prediction Model construction and Evaluation

Following systematic feature engineering and multicollinearity assessment, we employed binomial multivariable logistic regression to construct prediction models for prolonged LOS in frail older adults with multimorbidity. Logistic regression was selected for its clinical interpretability, transparent coefficient estimation, and established utility in healthcare prediction tasks, particularly in contexts where model explainability is paramount for clinical decision-making[19]. Model performance was assessed using Discrimination: Area Under the Receiver Operating Characteristic Curve (AUC-ROC). Calibration: Calibration plot, calibration slope, calibration-in-the-large, and Hosmer–Lemeshow goodness-of-fit test. Clinical Utility: Decision Curve Analysis (DCA) to evaluate net benefit across probability thresholds. Overall Accuracy: Sensitivity, specificity, positive predictive value (PPV), negative predictive value (NPV), and F1-score. Internal validation employed two complementary strategies:10-fold cross-validation, providing mean and standard deviation of performance metrics. Bootstrap validation (1,000 iterations) to estimate optimism and generate bias-corrected AUC values.

### Statistical Analysis

All analyses were performed using R (version 4.4.3; R Foundation for Statistical Computing, Vienna, Austria). Continuous variables were summarized as mean ± standard deviation (SD) when approximately normally distributed, or as median with interquartile range (IQR: 25th–75th percentiles) otherwise. Group comparisons for continuous outcomes were conducted using the Mann–Whitney U test, while categorical variables reported as counts and percentages were compared using Pearson’s chi-square (χ²) test. Predictive modelling, including predictor selection, model fitting, comprehensive internal validation and model performance was implemented entirely in R. Decision curve analysis was used to evaluate clinical utility across risk thresholds. A two-sided p-value < 0.05 was considered statistically significant.

## Results

A total of 597 hospitalized older adults with multimorbidity were screened between January 2022 and December 2023. After excluding 97 individuals who did not meet frailty criteria (Frailty Index <0.25) and 23 with incomplete records, 440 patients were included in the final analysis. Using stratified random sampling (seed = 123), participants were divided into a training set (n = 308, 70%) for model development and a test set (n = 132, 30%) for validation.

**Figure 1.**
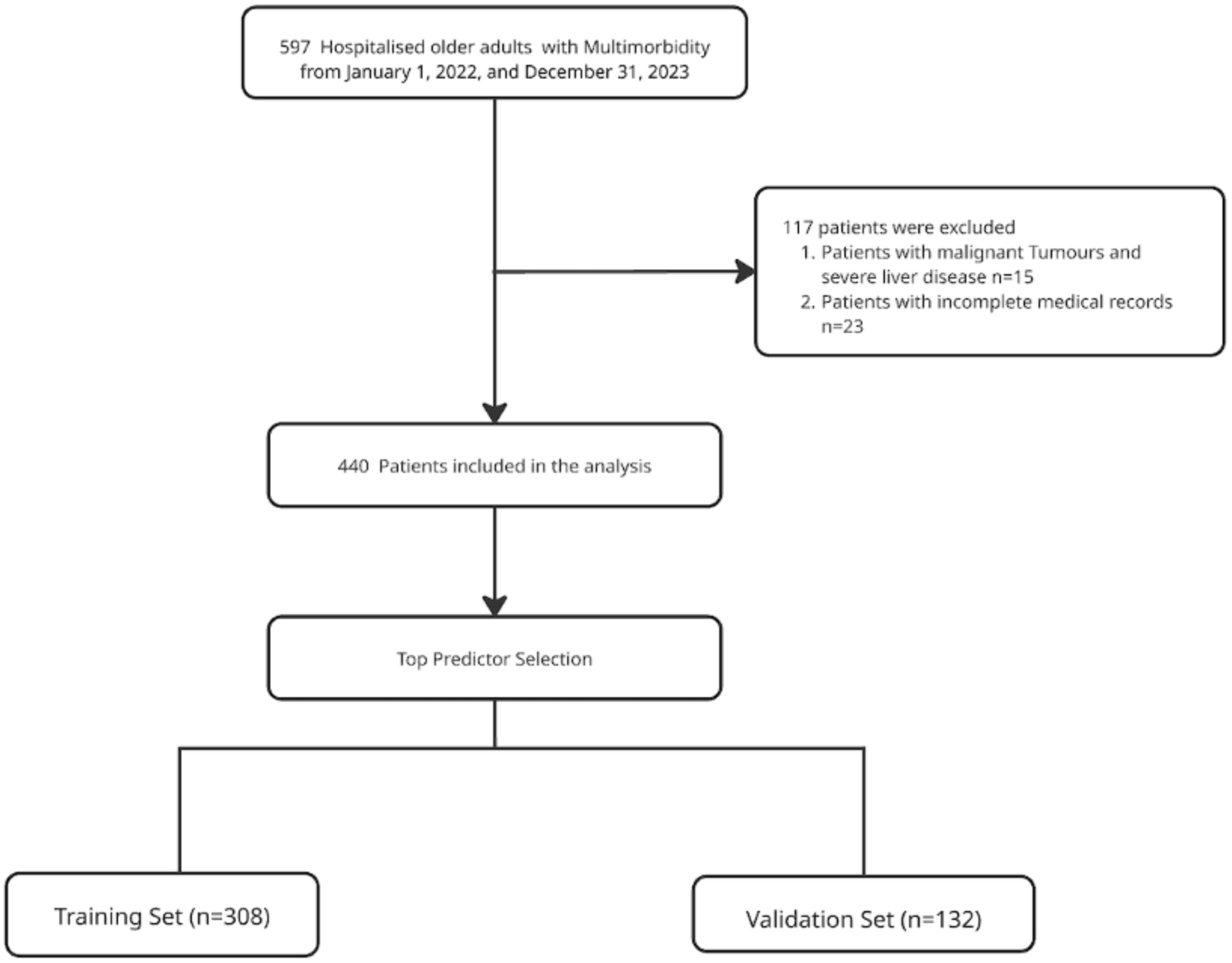
Study flowchart illustrating the inclusion and exclusion process,

### Baseline Clinical Characteristics

The median age of the cohort was 79 years (IQR: 71–87), and 51.8% were male. Comorbid hypertension (81.1%) and diabetes mellitus (49.1%) were the most prevalent chronic conditions, followed by prior stroke (47.5%). Median systolic blood pressure was 138 mmHg (IQR: 124–151), median serum albumin was 36.1 g/L (IQR: 31.9–40.0), and the median neutrophil-to-lymphocyte ratio (NLR) was 3.55 (IQR: 2.21–5.93). No significant differences were observed between training and test sets for any demographic or laboratory parameters (all p > 0.05), confirming that random stratification achieved balanced groups (Table 1).

| Variable | Total<br>N=440 | Derivation set<br>N=308 | Validation<br>N=132 | p value |
| --- | --- | --- | --- | --- |
| **Demographics** |  |  |  |  |
| Male, N (%) | 227 (51.8) | 163 (52.9) | 64(48.5) | **0.393** |
| Female, N (%) | 213 (48.2) | 145 (47.1) | 68(51.5) |  |
| Age, years (median [IQR]) | 79 (71, 87) | 80 (71, 89) | 79(71,85) | **0.287** |
| **Past Medical History** |  |  |  |  |
| Smoking, N (%) |  |  |  | **0.593** |
| Yes | 93 (21.1) | 63 (20.5) | 30 (22.7) |  |
| No | 347 (78.9) | 245 (79.5) | 102 (77.3) |  |
| Hypertension, N (%) |  |  |  | **0.770** |
| Yes | 357 (81.1) | 251 (81.5) | 106 (80.3) |  |
| No | 83 (18.9) | 57 (18.5) | 26 (19.7) |  |
| Diabetes, N (%) |  |  |  | **0.280** |
| Yes | 195 (44.3) | 126 (40.9) | 69 (52.3) |  |
| No | 295 (55.7) | 182 (59.1) | 63 (47.7) |  |
| Stroke, N (%) |  |  |  | **0.632** |
| Yes | 209 (52.5) | 144 (46.7) | 65 (49.2) |  |
| No | 231 (47.5) | 164 (53.3) | 67 (50.8) |  |
| **Patient Vitals** |  |  |  |  |
| Systolic Blood Pressure (median [IQR]) | 138 (123,151) | 138 (124,151) | 138 (121,151) | **0.954** |
| **Laboratory values** |  |  |  |  |
| Haemoglobin IU/L (median [IQR]) | 12.13 (10.36, 13.4) | 12.1 (10.34, 13.5) | 12.1(10.1,13.3) | **0.246** |
| Neutrophil, IU/L (median [IQR]) | 4.46 (3.3, 6.04) | 4.46 (3.46, 6.08) | 4.52 (2.97, 6.04) | **0.607** |
| N: L Ratio (median [IQR]) | 3.44 (2.19, 5.9) | 3.59 (2.26, 6.26) | 3.3 (2.09, 5.39) | **0.444** |
| TC, mmol/L (median [IQR]) | 4.18 (3.26, 5.06) | 4.21 (3.26, 5.07) | 4.17(3.32, 4.94) | **0.626** |
| LDL-C, mmol/L (median [IQR]) | 2.55 (1.79, 3.16) | 2.53 (1.78, 3.17) | 2.57 (1.8, 3.13) | **0.858** |
| HDL-C, mmol/L (median [IQR]) | 1.0 (0.78, 1.2) | 1.02 (0.78, 1.12) | 0.98 (0.78, 1.22) | **0.881** |
| Non-HDL-C, mmol/L (median [IQR]) | 3.2 (2.34, 3.87) | 3.18 (2.32, 3.91) | 3.21 (2.37, 3.83) | **0.787** |
| HCY, μmol/L (median [IQR]) | 15.84 (12.1, 22.6) | 15.74 (12.09, 22.63) | 16.1(12.0, 23.1) | **0.243** |
| Scr, μmol/L (median [IQR]) | 73.55 (59.3, 98.4) | 73.5 (61.3, 99.6) | 73.9(57.6, 97.3) | **0.663** |
| Albumin, μmol/L (median [IQR]) | 35.75 (32, 39.9) | 36.19 (31.9, 40) | 35.4(32, 39.2) | **0.285** |

N: L, Neutrophil Lymphocyte ratio; TC, Total Cholesterol; LDL-C, Low-density lipoprotein; HDL-C, High-density lipoprotein; Non-HDL-C, Non-High-density lipoprotein; HCY Homocysteine C; Scr, Serum creatinine

### Feature Selection and Model Predictors

Among 14 candidate variables, the ensemble feature selection approach (Elastic Net, XGBoost with SHAP, and Boruta) consistently identified eight predictors strongly associated with prolonged hospital stay (>8 days): Age, Diabetes mellitus, Hypertension, Prior stroke, Serum albumin, High-density lipoprotein cholesterol (HDL-C), Systolic blood pressure (SBP) and Neutrophil-to-lymphocyte ratio (NLR). (Figure 2-4)

**Figure 2.**
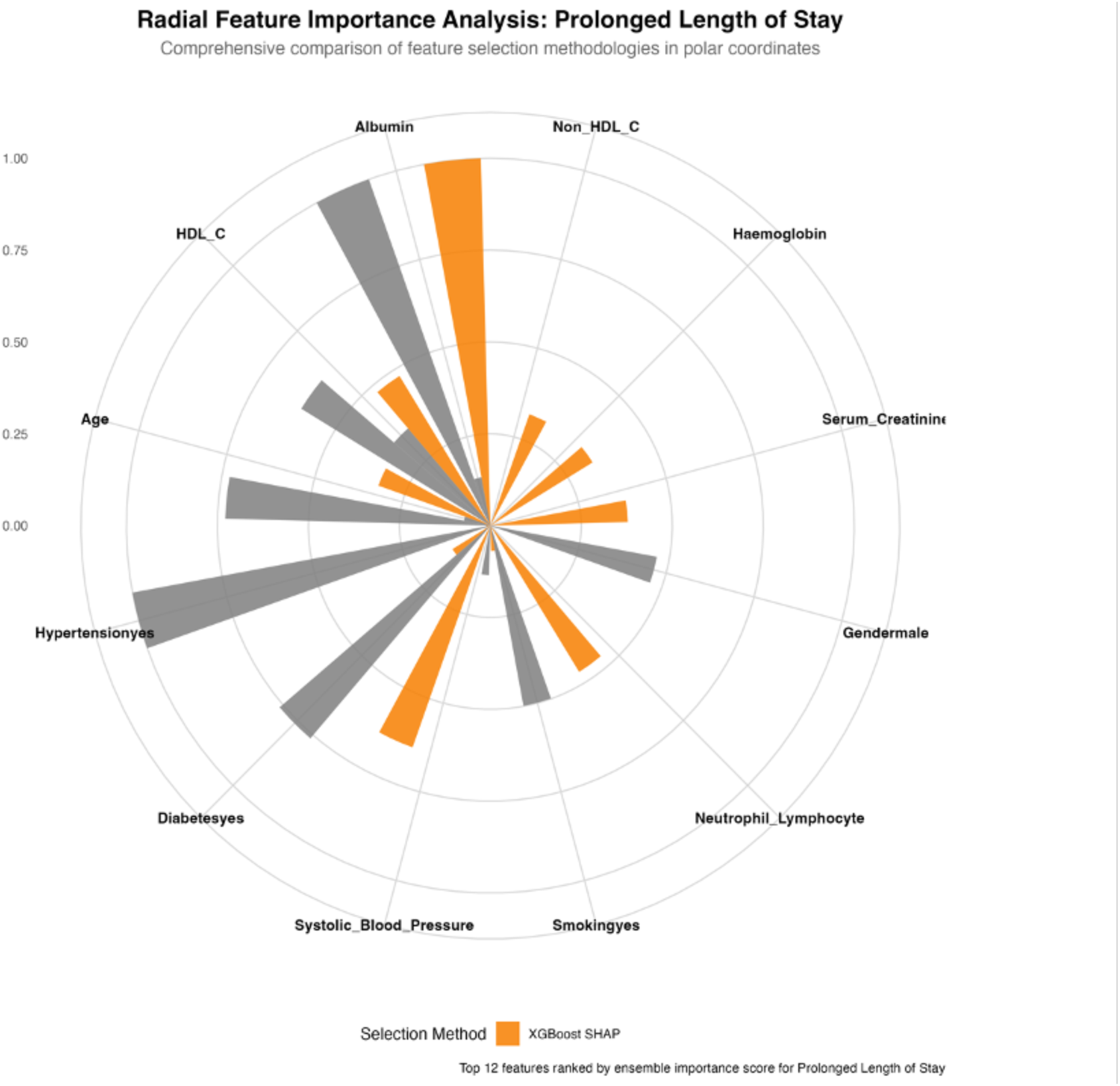
Radial Feature importance ranking based on SHAP (SHapley Additive exPlanations) values derived from the XGBoost algorithm

**Figure 3.**
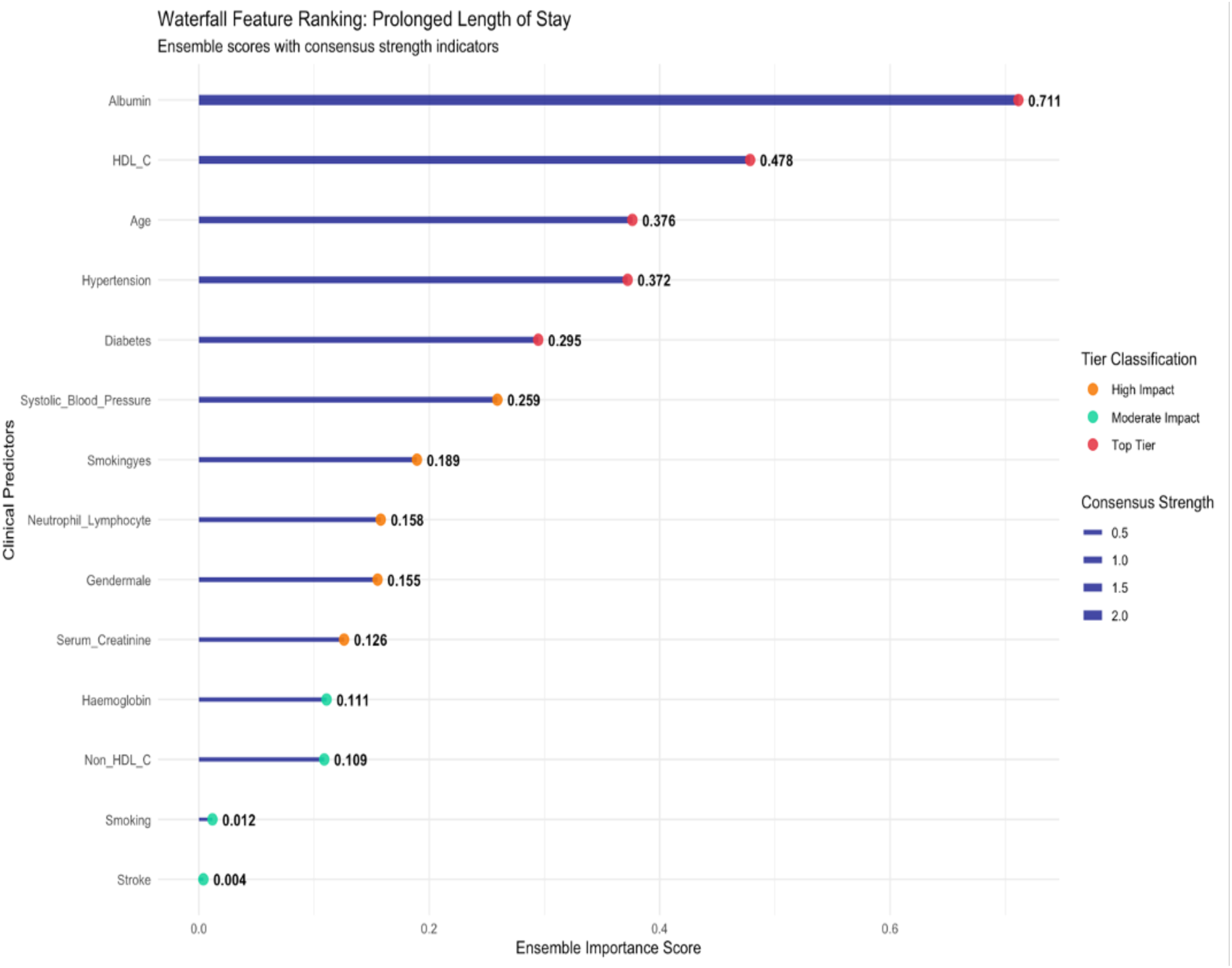
Waterfall presentation Integrated ensemble ranking of predictors across Elastic Net, SHAP, and Boruta algorithms

**Figure 4.**
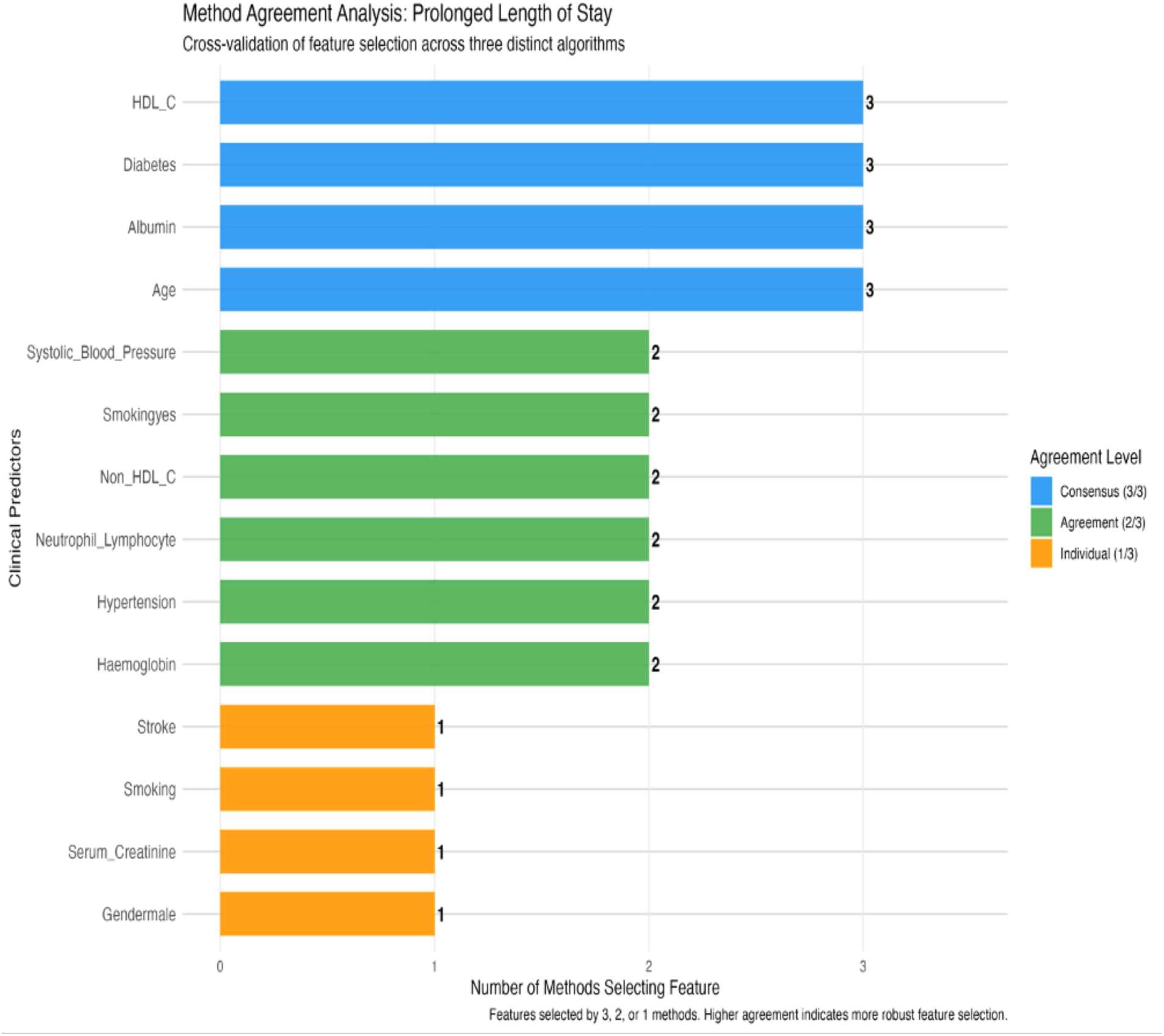
Method agreement analysis Integrated ensemble ranking of predictors across Elastic Net, SHAP, and Boruta algorithms

### Model Performance

The model demonstrated good discrimination in both training (AUROC 0.676, 95% CI 0.615-0.737) and test (AUROC 0.770, 95% CI 0.688-0.853) sets, demonstrating stable and generalizable predictive performance.

**Figure 5.**
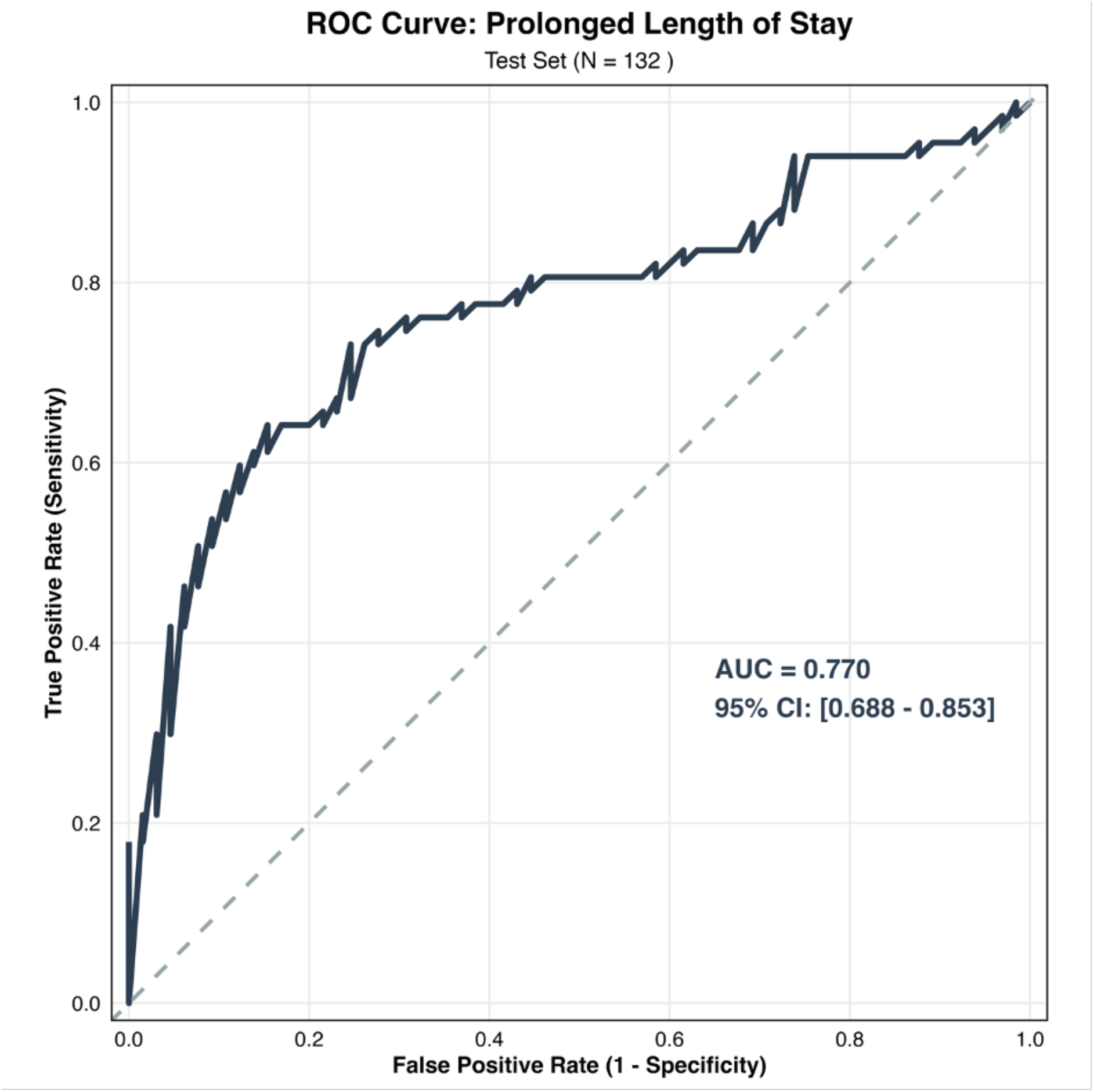
Receiver Operating Characteristic (ROC) curves for the predictive model in training and test datasets, demonstrating good discrimination

The Hosmer-Lemeshow goodness-of-fit test indicated acceptable calibration in the test set (χ² = 14.855, p = 0.062), suggesting that predicted probabilities aligned well with observed event rates across risk deciles. The model’s Brier scores were 0.222 (training) and 0.205 (test), indicating acceptable probabilistic accuracy. At the optimal Youden threshold (0.585), classification metrics were as shown in Table 2.

**Table 2.** Classification performance metrics for the predictive model in the test dataset.

|  |  |
| --- | --- |
| Accuracy | 77.1% |
| Sensitivity (recall) | 64.2% |
| Specificity | 84.6% |
| Positive predictive value (PPV): | 81.1% |
| Negative predictive value (NPV): | 69.6% |
| F1-score | 0.72 |

Internal validation confirmed model stability and low overfitting risk: 10-fold cross-validation: mean AUROC 0.687 (SD 0.072), mean accuracy 70.2% (SD 7.5%), mean sensitivity 76.4% (SD 16.3%), mean specificity 63.4% (SD 17.5%), mean Brier score 0.225 (SD 0.026) (Figure 6).

**Figure 6.**
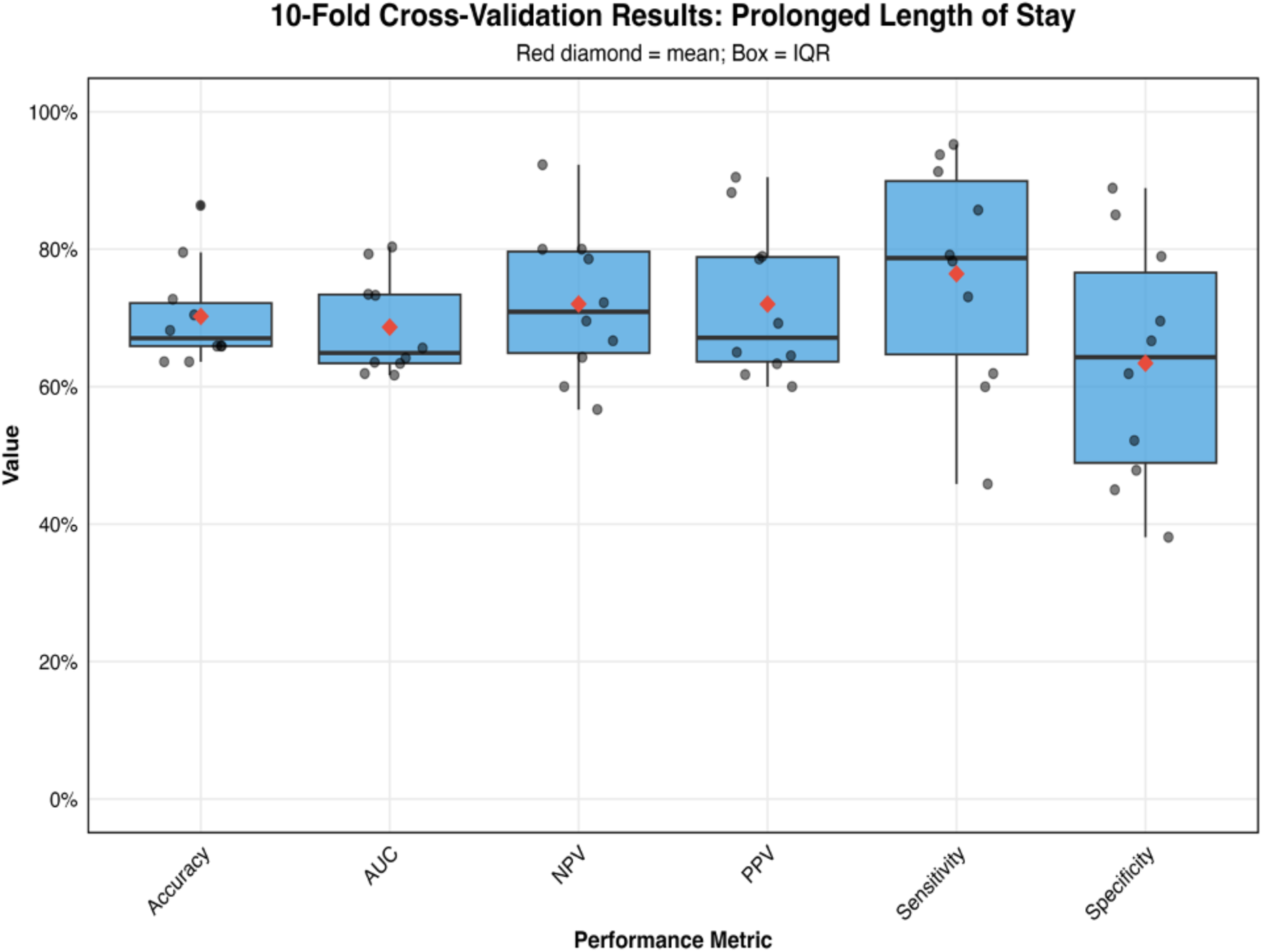
Box plot illustrating the distribution of six performance metrics across 10 folds of cross-validation for a predictive model designed to forecast prolonged hospital stays

Bootstrap validation (1,000 iterations): apparent AUC 0.729, out-of-bag AUC 0.685, resulting in a mean optimism of 0.044 (SD 0.052). The optimism-corrected AUC (0.672) closely aligned with both cross-validation and test-set results, confirming reproducibility. Decision curve analysis (DCA) demonstrated a consistent net clinical benefit across threshold probabilities of 0.05–0.60 compared with “treat all” and “treat none” strategies. The highest clinical benefit occurred at thresholds between 0.05–0.20, suggesting strong utility for guiding early, low-intensity interventions such as geriatric consultation or proactive discharge planning. (Figure 9).

**Figure 9:**
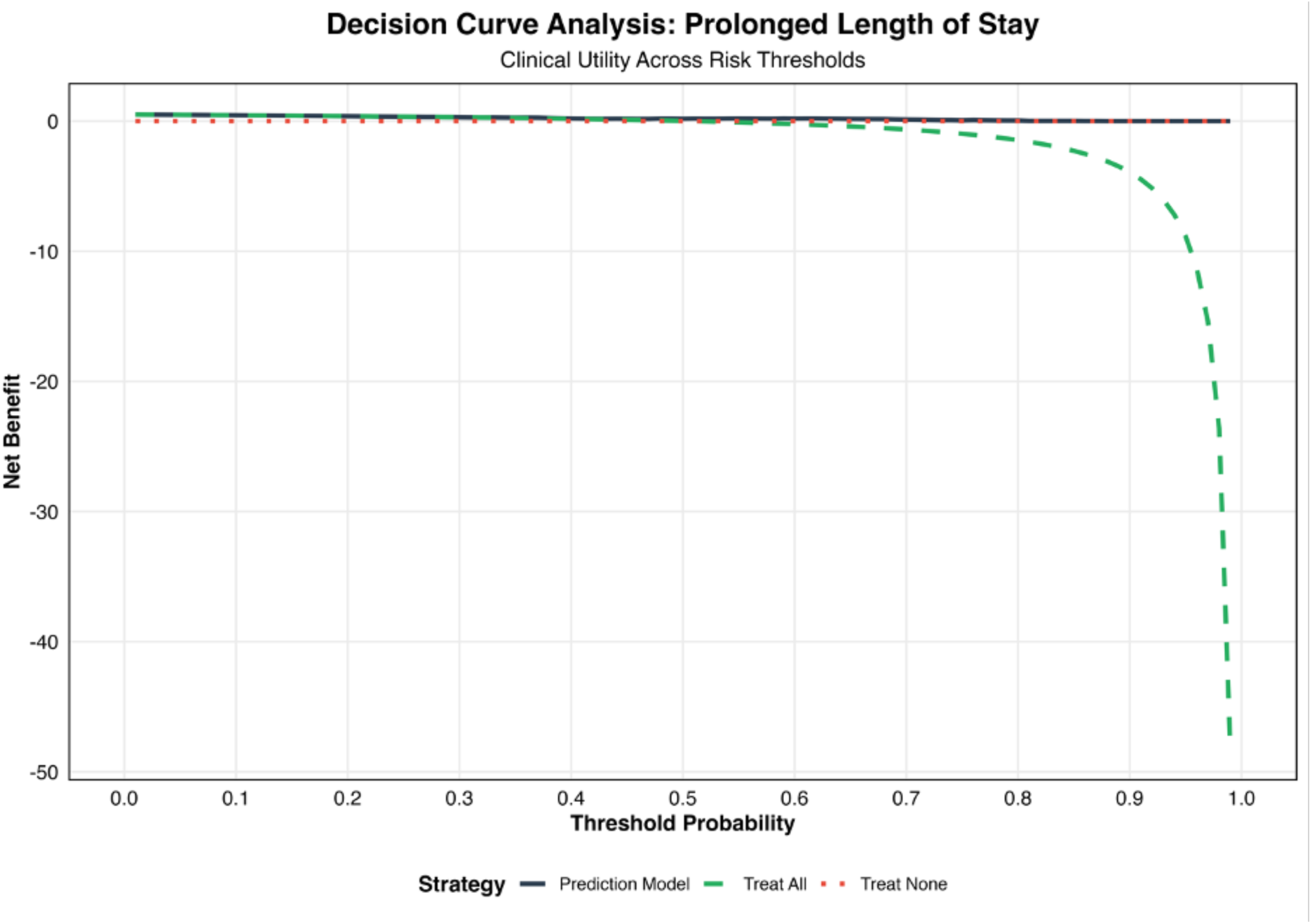
Decision curve analysis (DCA) depicting net clinical benefit across probability thresholds (0.05–0.60) compared with “treat-all” and “treat-none” strategies

The interpretable logistic regression model incorporating age, comorbidities, and simple laboratory markers reliably predicted prolonged LOS in frail older adults with multimorbidity. Performance was consistent across training, test, and cross-validation sets, with minimal optimism bias. Albumin emerged as the most protective biomarker, while elevated NLR and prior stroke were key predictors of adverse outcomes. The model demonstrates potential for integration into hospital information systems for real-time risk stratification.

### Clinical Implementation

The prediction model was implemented into a user-friendly clinical decision-support application. This digital tool integrates the model’s core predictors, comorbidities, demographic characteristics, and key laboratory biomarkers to provide real-time, individualized risk stratification at the point of care. The application delivers personalized risk estimates, evidence-informed clinical recommendations, comparative risk assessments across relevant outcomes, and generates exportable PDF reports designed for seamless integration into electronic health record systems, thereby facilitating clinical adoption and workflow integration (Figure 10).

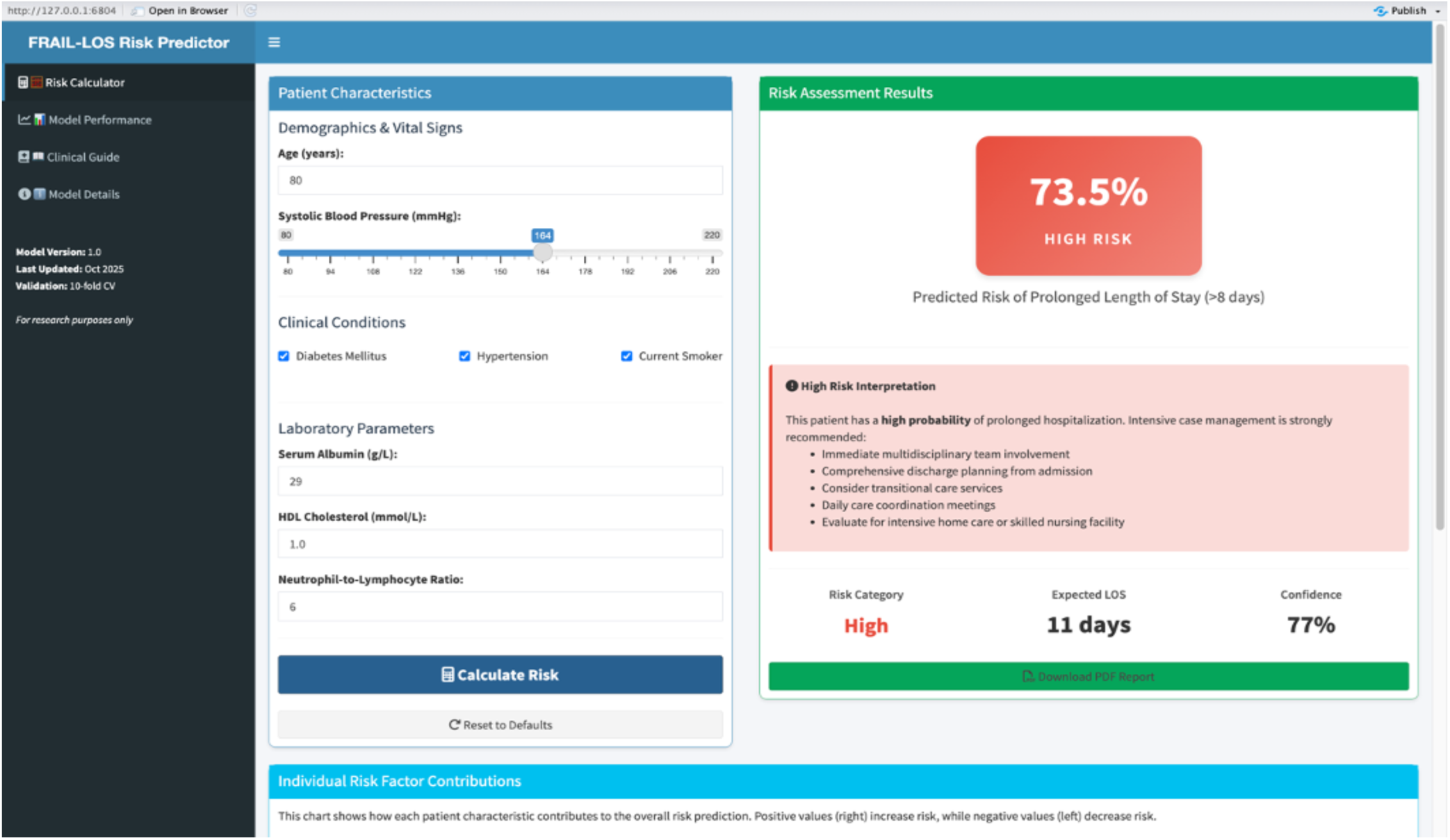

## Discussion

This study developed and internally validated an interpretable, machine learning–enhanced logistic regression model to predict prolonged hospital length of stay (LOS) among frail older adults with multimorbidity. Using routinely available demographic, clinical, and biochemical variables, the model achieved moderate-to-good discrimination (AUC = 0.770), acceptable calibration, and demonstrated consistent internal validity through cross-validation and bootstrap testing. These findings support the feasibility of integrating transparent, data-driven prediction models into geriatric inpatient care to guide individualized clinical decision-making. The relationship between frailty, multimorbidity, and adverse hospital outcomes has been well established, yet predictive tools tailored to this high-risk population remain limited. Previous models for length of stay have largely focused on general medical populations or have relied on non-geriatric predictors, often neglecting frailty-specific biological and functional dimensions. Empirical studies, including those by Lisk et al. (2019) and Bacchi et al. (2021), reported area under the curve (AUC) values ranging from 0.65 to 0.80 for their predictive models. However, these investigations did not explicitly address the interplay between frailty and multimorbidity, a critical confounder that may influence model performance and generalizability [21] [26]. Our model, which integrates simple yet physiologically meaningful indicators including albumin, HDL cholesterol, and the neutrophil-to-lymphocyte ratio (NLR) demonstrates that frailty-informed predictors can achieve comparable or superior performance while maintaining clinical interpretability. Several predictors identified in this study have strong biological plausibility in the context of frailty and aging. Serum albumin emerged as the most protective factor, aligning with extensive evidence linking hypoalbuminemia to inflammation, malnutrition, and poor recovery in older adults. Its inclusion underscores the importance of nutritional and inflammatory surveillance during hospitalization [20]. Elevated NLR was independently associated with prolonged LOS, consistent with research implicating chronic systemic inflammation as a core driver of frailty and delayed recovery. The ease of deriving NLR from routine blood counts enhances its value as a pragmatic biomarker in geriatric wards [21]. Low HDL cholesterol and higher systolic blood pressure were linked with adverse outcomes, possibly reflecting the vascular and metabolic dysregulation common in frail, multimorbid adults[22]. History of stroke and advanced age further compounded risk, illustrating the cumulative impact of vascular burden and physiological decline on recovery potential [23][24].These predictors, readily obtainable from routine clinical data, collectively reflect the multidimensional vulnerability characteristic of frailty a hallmark that traditional models rarely capture. To our knowledge, this is among the first studies to develop a transparent, internally validated predictive model for prolonged hospitalization specifically in frail, multimorbid older adults. The integration of multiple feature selection methods Elastic Net, XGBoost with SHAP, and Boruta ensures robustness and reduces model bias, while the use of logistic regression preserves interpretability crucial for clinical implementation. Unlike black-box algorithms that offer little insight into decision rationale, our model provides direct, interpretable coefficients and clinically meaningful odds ratios. This hybrid approach bridges the gap between conventional epidemiology and modern machine learning, embodying the growing paradigm of explainable artificial intelligence in geriatric medicine[25].

Furthermore, by demonstrating strong internal validation (10-fold cross-validation and 1,000 bootstrap resamples) and modest optimism, the study establishes a reproducible methodological foundation. The model’s decision curve analysis showed clear net benefit at low threshold probabilities (0.05–0.20), suggesting its suitability for guiding low-intensity, early interventions such as proactive discharge planning, nutritional optimization, and early geriatric consultation. This clinical applicability is particularly relevant for resource-constrained hospital systems managing complex older adults.

From a practical standpoint, the model can be integrated into electronic health record systems to automatically generate real-time risk scores upon admission. Such integration would enable clinicians to prioritize comprehensive geriatric assessments for high-risk patients and streamline discharge planning. In gerontology practice, the model supports precision geriatric medicine delivering interventions proportionate to an individual’s physiological resilience rather than chronological age. In public health and systems-level planning, these findings highlight the potential of interpretable AI tools to enhance efficiency, reduce avoidable bed-days, and inform policies on geriatric care resource allocation

## Limitations

Despite the strengths of this study, several limitations should be acknowledged. First, this was a single-centre study, which may limit generalizability to other healthcare settings and populations. External validation using multicentre and demographically diverse cohorts is necessary to confirm model robustness. Second, frailty was assessed cross-sectionally at admission; temporal changes in frailty status or functional recovery trajectories were not captured. Future work integrating longitudinal frailty measures may improve prediction of dynamic outcomes such as readmission. Third, while missing data were handled using multiple imputation, residual confounding inherent in retrospective designs cannot be excluded. Lastly, although the model demonstrated strong internal validity, real-world implementation studies are needed to assess its clinical impact, usability, and potential for bias mitigation across subgroups.

## Conclusion

This study presents a clinically interpretable, machine learning–informed prediction model capable of identifying multimorbid frail older adults at risk for prolonged hospitalization using readily available admission data. By balancing predictive accuracy with transparency, the model addresses a key gap in geriatric prognostication and offers a scalable solution for precision risk stratification in aging care. Clinically, this model can support precision risk assessment at admission, enabling proactive discharge planning, early geriatric consultation, and more efficient allocation of hospital resources. Theoretically, it advances the emerging field of explainable artificial intelligence in gerontology demonstrating that data-driven tools can enhance care for older adults without sacrificing interpretability. The findings advance the intersection of clinical epidemiology, gerontology, and artificial intelligence, contributing to the ongoing effort to make hospital care for older adults more anticipatory, individualized, and efficient.

## Data availability

The data underlying this article will be shared upon request to the corresponding author.

